# Influence of Lamina Vastoadductoria Dissection on the Outcomes of Femoral Artery Extensive Lesion Stenting: A Pilot Randomised Investigation

**DOI:** 10.1101/2022.02.24.22271436

**Authors:** Andrey A. Karpenko, Artem A. Rabtsun, Irina V. Popova, Shoraan B. Saaya, Alexandr A. Gostev, Pavel V. Ignatenko, Vladimir B. Starodubtsev, Alexey V. Cheban

## Abstract

**Objective:** Aim of the study was to improve the immediate and long-term results of stenting of the superficial femoral artery in extended lesions with the changing of the biomechanics of SFA and of the first portion of the popliteal artery.

**Methods:** Pilot randomized prospective single-center study were included 70 patients. Patients were randomized into two groups in 1×1 format for 35 people using the envelope method. Self-expanding bare metal stents were used in all cases. At the first group standard revascularization procedures with SFA stenting were performed; in the second group, the superficial femoral artery stenting was supplemented with fasciotomy in the Hunter’s canal with the superficial femoral artery intersection. The total observation period was 2 years. During the observation period an assessment of the clinical symptoms of the lower extremities, measurement of the ankle-brachial index and ultrasound duplex scanning of the operated segment were performed.

**Results:** All procedures in both groups were successfully performed. Primary patency through 24 months was 28.5% (10 of 35) in group 1 and 60% (21 of 35) in group 2 (p=0,015).

**Conclusions:** Changing the biomechanical properties of the distal of the superficial femoral artery segment and of the first portion of the popliteal artery is safe and contributes to the primary patency improvement during the stenting of extended of the superficial femoral artery lesions compared to standard SFA stenting. Dissection of the lamina vastoadductoria with transection of the collateral branches of the knee joint network reduces frequent and severe damages of stents after the stenting of the superficial femoral artery extended lesion.

According to the frequency of complications in the early and mid-term postoperative period, limb salvage, mortality and the secondary patency rates, the new method is comparable with standard of the superficial femoral artery stenting.

## Introduction

When comparing the effectiveness of open and endovascular interventions for superficial femoral artery (SFA) extended lesions in 1 year, the primary patency rates are comparable and range from 50% to 90%.^1-7^ In the 3-year follow-up period, the primary patency rates for bypass surgery were 73% versus 42% for the stented segment (hazard ratio [HR], .4; 95% confidence interval, .23-.71). Endovascular interventions have lower rates of mortality and complications both in the early and long-term follow-up.^8^

Among complications of SFA extended lesions treatment, restenosis had a high occurrence during a year follow-up (more than 60%).^6,9-13^ Moreover, fractures of implanted stents reached 50% of the total implantations.^14^ The frequent combination of restenosis of the operated arterial segment and fractures of implanted stents contributed to further study and changes in the mechanical properties of nitinol stents and physiological deformities of the operated artery during knee joint movement.^14-16^ However, a significant risk of fractures remains even for second-generation stents, which increases the rate of restenosis and reocclusion to approximately 37% within a year.^17^

Many authors note a relationship between the development of restenosis and variable biomechanics of SFA and the first portion of the popliteal artery. The main trends for improving long-term results of interventions aimed at changing the physical properties of devices, although there were no studies aimed at changing the biomechanical properties of the SFA and the first portion of the popliteal artery. Considering this, the search for an optimal method of SFA extended lesion treatment is currently one of the main investigations in vascular surgery.^18,19^

This study aimed to improve the immediate and long-term results of SFA stenting in extended lesions with the changing of the biomechanics of SFA and the first portion of the popliteal artery.

## Materials and Methods

This pilot randomised prospective single-centre study was approved by the Local Ethics Committee of Meshalkin National Medical Research Centre, Ministry of Public Health of the Russian Federation (September 15, 2015, protocol No. 53). All patients were included in the study after obtaining their informed consent. We included 70 patients with TASC II D lesions of SFA from September 2015 to August 2017. Inclusion and exclusion criteria are presented in Table 1. In addition, patients with patent 2 and 3 segments of the popliteal artery were selected.

**Table 1.** Criteria for inclusion and exclusion.

| Inclusion criteria | Exclusion criteria |
| --- | --- |
| 45-80 years old | Chronic heart failure III -IV functional class according to NYHA classification |
| Primary lesion of the superficial femoral artery of class D according to TASC II | Chronic decompensated “pulmonary” Heart |
| Ischemia of the lower extremity 3-6 c according to Rutherford | Severe hepatic or renal failure (bilirubin > 35 mmol/L, glomerular filtration rate < 60 mL/min) |
| Consent to participate in the study | Polyvalent drug allergy |
| Satisfactory flow of outflow | Malignant oncological diseases in the terminal stage with a projected lifespan of up to 6 months |
|  | Acute cerebrovascular accident |
|  | Severe calcification of the arteries of the lower extremities tolerant to balloon angioplasty |
|  | Patients with significant lesions of the common femoral artery |
|  | Refusal of the patient to participate or to continue participation in the study |

NYHA, New York Heart Association; TASC, Trans-Atlantic Inter-Society Consensus

Patients were randomised into two groups in 1×1 format for 35 people using the envelope method. (Figure 1). Characteristics of groups in the table 2. In the first group, standard revascularisation procedures with SFA stenting were performed; in the second group, SFA stenting was supplemented with fasciotomy in Hunter’s canal with SFA intersection. At the hospital stage, the hemodynamic characteristics of all patients were assessed by measuring the ankle-brachial index and duplex scanning of lower limb arteries on the device VOLUSON 730 (GE Healthcare, Zipf, Austria). Multidetector computed tomography (MDCT) angiography of the lower limb arteries was also performed on a 320-slice computed tomography scanner Aquilion One (Toshiba, Tokyo, Japan), and a contrast agent Iomeron 400 (Bracco, Milan, Italy) was used to clarify anatomical features and volume of the lesion of the arteries. Drug therapy included the administration of acetylsalicylic acid before the procedure (300 mg/day), starting at least a day. After the procedure, all patients received acetylsalicylic acid (100 mg per day) for a lifetime and clopidogrel (75 mg per day) for 3 months. Access to the arteries by ipsilateral or contralateral puncture of the common femoral artery was performed. Recanalisation with a hydrophilic 0.035-inch guide was performed. Anticoagulant (heparin sulfate [150 U/kg] was introduced before starting the procedure. After angioplasty, a nitinol self-expanding bare metal stent was implanted in accordance with the recommendations of the American College of Cardiology/American Heart Association.^20^ If necessary, several stents were implanted (maximum 2). Stent implantation was not performed in the middle or distal third of the popliteal artery. Drug-eluting devices were not used in our study. After stenting of SFA in the second group under local anaesthesia, access was made in the middle third of the thigh on the inner surface in the projection of Hunter’s canal, and the sartorius muscle was moved down. Subsequently, intermuscular vastoadductor sept was dissected, a n d SFA was mobilised throughout. The arteries that fixate SFA were ligated and dissected. The total observational period was 2 years. During the observation period, an assessment of clinical symptoms of the lower extremities, measurement of the ankle-brachial index (ABI), and ultrasound duplex scanning of the operated segment were performed. When stenosis or occlusion was detected during the observation period, confirmed by ultrasound, the patients underwent MDCT-angiography of the lower limb arteries.

**Figure 1.**
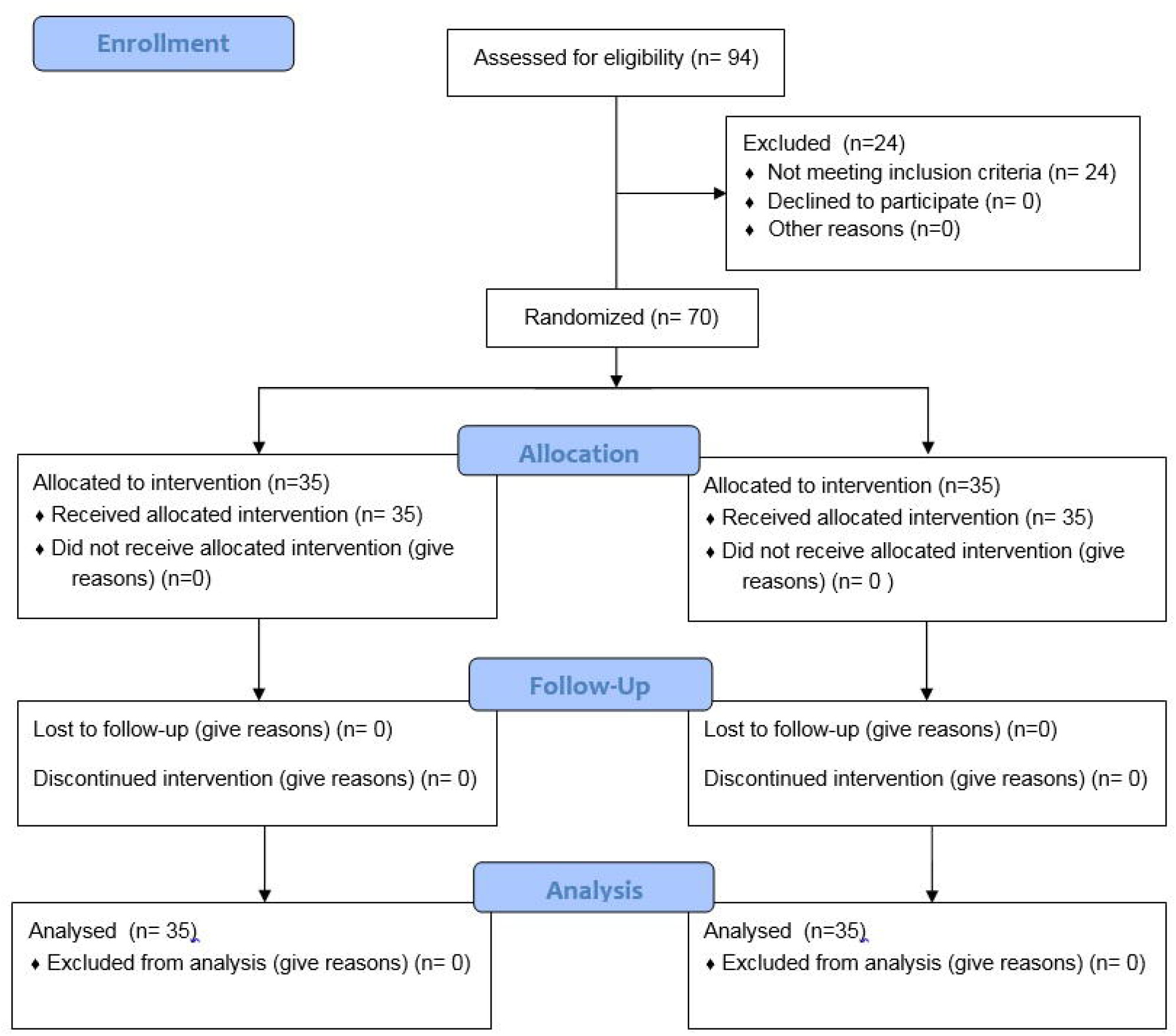
Patient inclusion and randomisation scheme (CONSORT)

**Table 2.** Characteristics of groups.

|  | **Group 1 (n = 35)**<br>**(Stenting SFA)** | **Group 2 (n = 35)**<br>**(Stenting SFA**<br>**+fasciotomy)** | **p** |
| --- | --- | --- | --- |
| **Sex M/F** | 25/10 | 24/11 | N/A |
| **Average age (±SD)** | 65±6.62 | 66.7±9.39 | .256 |
| **Medium ABI (±SD)** | 0.49±0.114 | 0.51±0.096 | 1 |
| **Arterial hypertension** | 31 (89%) | 32 (91%) | .671 |
| **Hypercholesterolemia** | 14 (40%) | 20 (57%) | .396 |
| **Smoking** | 17 (49%) | 22 (63%) | .393 |
| **Chronic renal failure** | 13 (37%) | 8 (23%) | .196 |
| **Diabetes mellitus** | 13 (37%) | 6 (17%) | .538 |
| **Coronary heart disease** | 28 (80%) | 28 (80%) | 1 |

SD, standard deviation; ABI, ankle-brachial index; M, male; F, female; SFA, superficial femoral artery

### Primary endpoint

The primary safety endpoint was violation of limb function of the proposed method compared with standard endovascular recanalisation and the SFA stenting (i.e. Will dissection of the fascia of the adduction canal limit the patient’s everyday activities?). The primary efficiency endpoints were the primary vessel patency, which was rated as the absence of binary restenosis (50% or more) and reocclusion.

### Secondary composite endpoint

Stent fractures, limb salvage, secondary patency of the operated segment, and intraoperative complications were the secondary composite endpoint.

## Results

The distribution of the limb ischemia in groups and characteristics of arterial lesions is shown in Table 3.

**Table 3.** The distribution of limb ischemia in the groups and characteristics of arterial lesions.

| **Limb ischemia degree**<br>**(Rutherford)** | **Group 1**<br>**(Stenting**<br>**SFA)**<br>**(n = 35)** | **Group 2**<br>**(Stenting SFA**<br>**+fasciotomy)**<br>**(n = 35)** | **p** |
| --- | --- | --- | --- |
| **3** | 29 (83%) | 25 (71%) | .32 |
| **4** | 4 (11%) | 7 (20%) | .41 |
| **5** | 1 (3%) | 3 (9%) | .61 |
| **6** | 1 (3%) | 0 | 1 |
| **Characteristics of the lesion of the arteries** |  |  |  |
| **Stenosis / Occlusion** | 12/23 | 10/25 | .741 |
| **The length of the**<br>**lesion of the arteries**<br>**(±SD)** | 22.92 ± 5.62<br>(mm) | 21.2 ± 5.42<br>(mm) | 1 |
| **Average artery**<br>**diameter (±SD)** | 5 ± 0.81 (mm) | 4.73 ± 0.76<br>(mm) | .15 |
| **Outflow vessel by**<br>**Rutherford** | 6.32 ± 1.71<br>(points) | 6.04 ± 2.43<br>(points) | .79 |

SD, standard deviation; SFA, superficial femoral artery

Arterial lesions were localised only within SFA and the first portion of the popliteal artery in all patients. The average lesion length was 22.92 ± 5.62 cm in the first group and 21.2 ± 5.42 in the second group, (p = 1). The average diameter of SFA was 5 ± 0.81 mm in the first group and 4.73 ± 0.76 mm in the second group, (p = .15). The diameter of SFA was measured at the border of the distal third of SFA and the first portion of the popliteal artery. Total occlusions were 23 (65.7%) and 25 (71.4%) in the first and second groups, respectively (p = .741). Stenosis were 12 (34.3%) and 10 (28.6%) in the first and second groups, respectively (p = .741). The outflow vessel, assessed by Rutherford, was comparable in both groups. It was 6.32 ± 1.71 points in the first group and 6.04 ± 2.43 points in the second group with p = .79. In most cases, the implantation of a single stent was required to eliminate the artery lesion; however, based on the extent of the arterial lesion, the use of several stents was required (maximum of 2). There were no mortality cases during hospitalisation, 30-day follow-up period after discharge, and 2-year observational period. During the observational period, one amputation was performed in each group. In the first group, one amputation at the hip level was carried out at 6 months of observation, while the patient with diabetes, and limb ischemia initially corresponded to 5 stages by Rutherford. In the second group, a below-knee amputation was performed at 12 months, which is probably related to the distal embolism in the tibial artery with the development of acute ischemia. In the postoperative period, an increase in ABI was observed in both groups. The dynamics of the change of ABI in the control points between the groups did not differ significantly over all periods of observation. One patient dropped out of each group during the entire observational period. These are patients who underwent limb amputation.

The prediction of the development of restenoses/reocclusions from comorbidities was diabetes mellitus. When assessing the impact of the type of surgical treatment using logistic regression on the development of restenosis/reocclusion, superiority was found in patients with fasciotomy. According to the results of the odds ratio, the chances of a significant difference between the groups based on surgical treatment were not identified (Table 4).

**Table 4.** Predictors of restenosis and reocclusion of the operated segment (odds ratio and risk ratio).

|  | Odds ratio (CI 95%) |  | Risk ratio (CI 95%) |  |
| --- | --- | --- | --- | --- |
| Predictor |  | p |  | p |
| Arterial hypertension | .69 (.1; 4.8) | .7 | 1.32 (.31; 5.6) | .71 |
| Hypercholesterolemia | .44 (-1.98; .34) | .16 | 1.52 (.66; 3.47) | .32 |
| Smoking | .84 (.23; 3.13) | .79 | .99 (.44; 2.21) | .98 |
| Chronic renal failure | .74 (-1.5; .93) | .62 | 1.07 (.46; 2.5) | .87 |
| Diabetes | .18 (.04; .81) | .02 | 2.5 (1.1; 5.65) | .02 |
| Coronary heart disease | 1.1 (.26; 4.58) | .89 | .98 (.37; 2.62) | .97 |
| Treatment method | 3.78 (1.1; 12.5) | .026 | .47 (.2; 1.09) | .08 |
| Outflow vessel | .94 (.77; 1.15) | .56 | 1.04 (.79; 1.37) | .75 |

CI, confidence interval; p, p-value

There was a moderate correlation between SFA diameter and primary patency (R = -0.32, p =.03). The degree of ischemia did not affect the primary patency over the entire observational period in our study. Primary patency for the first group was: 3 months, 72%; 6 months, 60%; 12 months, 36%; and 24 months, 28.5%. For the second group, the primary patency was: 3 months, 80%, 6 months, 76%, 12 months, 72%; and 24 months, 60% (p = .02) (Figure 2).

**Figure 2.**
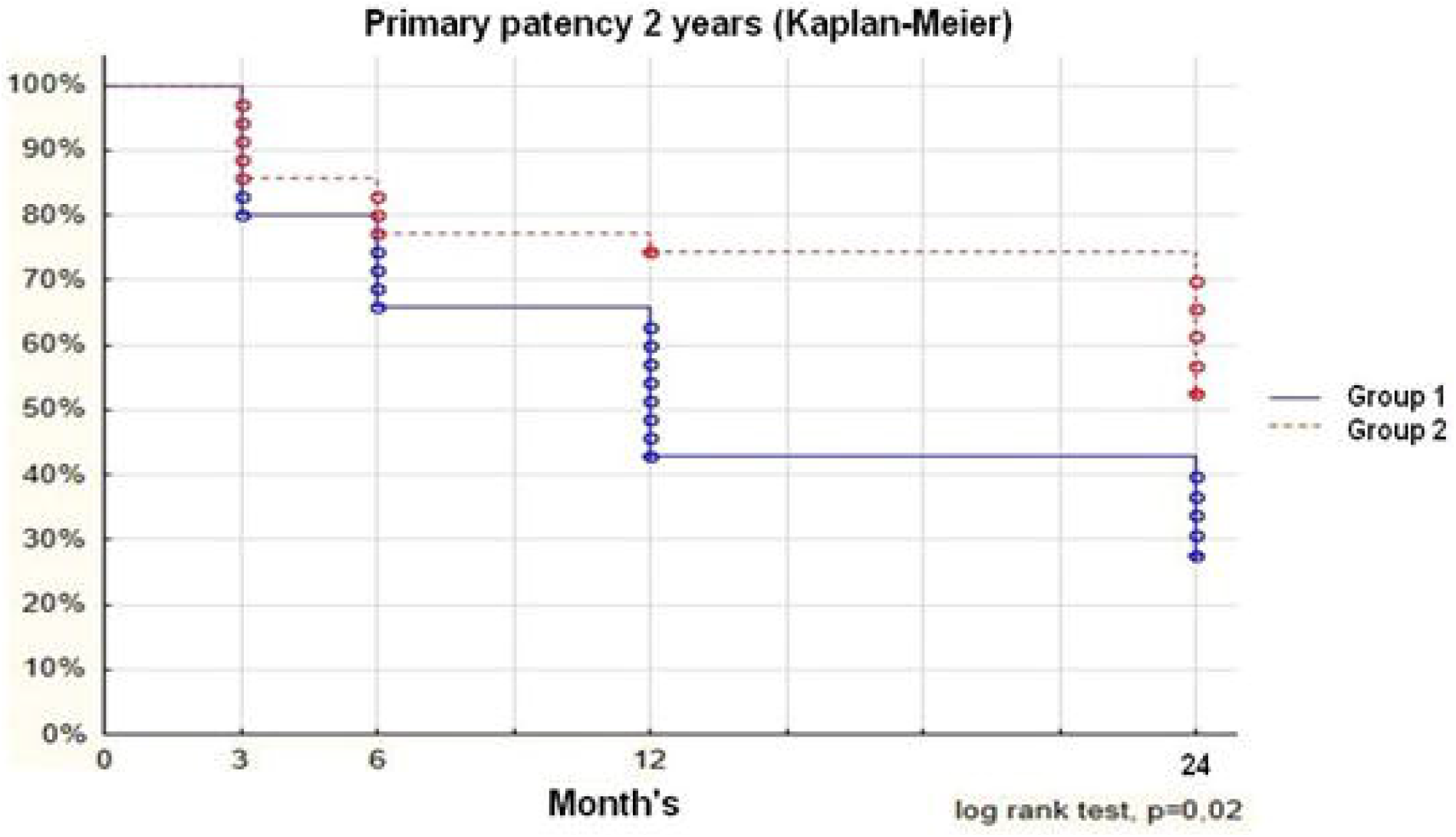
Primary patency of the operated segment.

The degree of stent fracture was assessed by radiography of the stented area in the frontal and lateral projections. The severity of stent fracture was assessed using a stent fracture grading modified from Rocha-Singh.^14^ We used the Mann-Whitney criterion to assess the frequency and severity of stent fracture between the groups (Figure 3). Based on the severity, the distribution in the groups was as follows: Group 1 (SFA stenting): type 0, 14%; type 1, 20%; type 2, 20%; type 3, 26%; type 4, 11%; type 5, 9%; and Group 2 (SFA stenting + fasciotomy): type 0, 37%; type 1, 23%; type 2, 14%; type 3, 11%; type 4, 9%; type 5, 6%. According to the results, the frequency and severity of stent fractures were significantly lower in the second group (p = .03). However, when assessing the effect of the stent fractures on the permeability of the stented segment using the logistic regression method, no significant effect was found (.78 [.56; 1.0] p = .11). There was no significant difference between the groups by calculating the combined secondary endpoint. Freedom from the development of secondary composite endpoints was four cases (11.5%) and eight cases (22.9%) in the first and second groups, respectively (p = .34). During the observational period, all patients received antiplatelet therapy as prescribed. In patients with significant restenosis/reocclusion of the operated segment, acute ischemia cases did not occur, although there was a return of chronic limb ischemia to the initial level. In patients with ischemia 4-6 degrees according to Rutherford, ischemia returns to the 3^rd^ stage. The analysis of the degree of ischemia included 34 patients in each group. The distribution of the degree of limb ischemia by category according to Rutherford after 24 months was as follows: Group 1 (SFA stenting)/Group 2 (SFA stenting + fasciotomy) accordingly: stage 0, 10 (29.41%)/15 (44.12%) (p = .16); stage 1, 8 (23.53%)/6 (17.65%) (p = .38); stage 2, 10 (29.1%)/9 (26.47%) (p = .5); and stage 3, 6 (17.65%)/4 (11.76%) (p = .37). The secondary patency was 60% in the first group and 57.1% in the second group. The ratio of stenoses and occlusions was in Group 1 (SFA stenting): 9 stenosis and 16 occlusions, in Group 2 (SFA stenting + fasciotomy): 5 stenoses and 9 occlusions. By the range of repeated interventions in Group 1 (SFA stenting): balloon angioplasty one case, balloon angioplasty with stenting two cases, below knee femoral-popliteal bypass with autovein in-situ three cases, and one case of below knee femoral-popliteal bypass with reversed autovein. In Group 2 (SFA stenting + fasciotomy) distribution was as follows: balloon angioplasty two cases, balloon angioplasty with stenting one case, and below knee femoral-popliteal bypass with polytetrafluoroethylene graft two cases. There was no significant difference between the groups.

**Figure 3.**
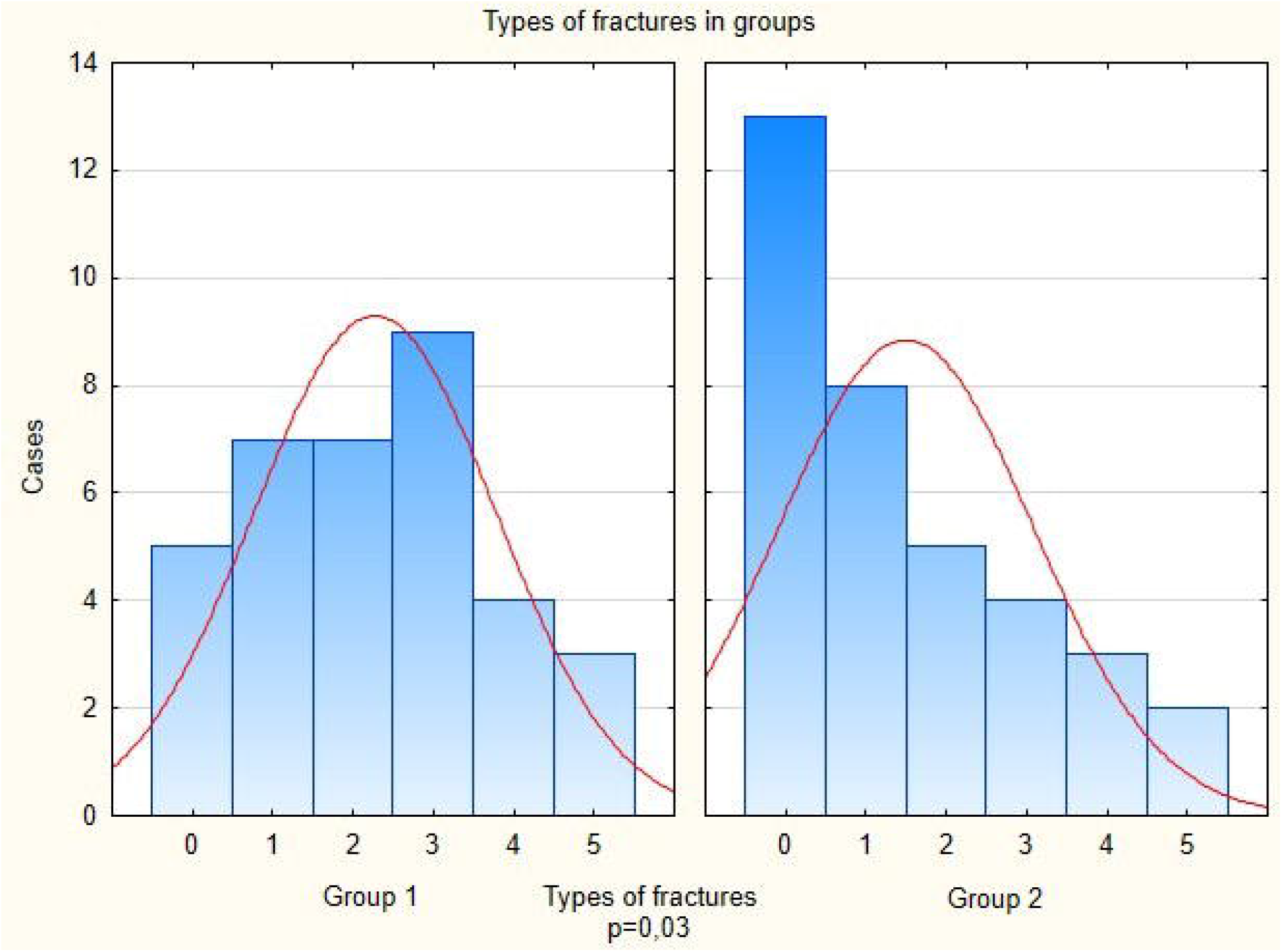
Frequency and severity of stent fractures between the groups.

## Discussion

With the increasing life expectancy of the population, the incidence of peripheral artery atherosclerosis also increases. Trends in reconstructive interventions on the arterial vessel in various basins aimed at increasing the proportion of endovascular procedures, as this contributes to less trauma, which is important for the age group of patients with several comorbidities. The treatment strategy for SFA lesions has changed in recent years. Endovascular techniques have shown an obvious advantage in the treatment of SFA classes A and B (TASC II). Stenting technology in the treatment of short and extensive SFA lesions showed an advantage over balloon angioplasty.^14-16,21^ Conversely, the use of balloons and stents with drug coating reduces the intensity of neointimal hyperplasia.^16,17,22^ While endovascular surgery is in the first place in the treatment of atherosclerotic lesions of SFA classes A and B, discussions continue for extended lesions of classes C and D. Despite the high percentage of technical intraoperative success, long-term results according to the literature remain unsatisfactory.^5, 6^

When comparing balloon angioplasty with stenting for extensive lesions of SFA, stenting has the best indices of primary patency in a short-term period.^15,19^ However, some studies have shown that with an increase in the length of the stented segment, the risk of stent fractures also increases.^6, 21^ To a certain extent, this is due to the unique features of SFA biomechanics, which are not found in other vessels.^23^ This study aimed to determine the effect of the new technique on the results of stenting of SFA extended lesions (TASC-II D) with the use of self-expanding nitinol bare-metal stents. Our results on primary patency within the 2-year follow-up in patients with fascia dissection was 60%, which is comparable and higher than what is observed in retrospective studies such as DURABILITY-200 (Physician Initiated Trial Investigating the Efficacy of the Implant of EverFlex 200mm Long Nitinol Stents in TASC C&D Femoropopliteal Lesions) (64.8%), STELLA (Long Superficial Femoral Artery Stenting With SuperA Interwoven Nitinol Stents) (66%), SUPERStudy (Randomised Trial of the SMART Stent versus Balloon Angioplasty in Long Superficial Femoral Artery Lesions) (45.9%). Simultaneously, in the DURABILITY-200 study, in 51% of cases, the length of the SFA lesion varied from 160 mm to 450 mm, and in 41% of cases from 150 mm to 200 mm. In the DURABILITY-200, the outflow vessel was represented by all three tibial arteries in 75% of cases, and only one tibial artery functioned in 9% of patients. Concerning the SUPERStudy, the lesion length ranged from 50 mm to 220 mm, and the average lesion length in the groups was 123 and 116.8 mm. In the SUPERStudy, the lesions of SFA more than 150 mm in the studied groups were 28.4% and 25.0%. The outflow vessel was not evaluated in the SUPERStudy. According to the results of the Supera 500 register, the average length of the lesion was 126 mm, and occlusions were detected in 53% of cases. In our study, the primary patency was 75% for 24 months. When compared with the control group, the primary permeability was significantly higher (28.5% vs. 60%). Thus, in the studies analysed, the extent of the SFA lesion varies significantly and only a few patients have extended lesions. None of the studies analysed the primary patency in subgroups according to the extent of the lesion. Only one study described the distribution of patients along the outflow vessel, of which most patients had a good peripheral vessel. The length of the affected segment in our study is comparable between groups (22.92 ± 5.62 cm and 21.2 ± 5.42 cm), and with other studies: 24.2 cm (DURABILITY-200) and 22 cm (STELLA). Contrarily, our study included patients with extensive lesions of SFA, which affected the low primary patency in the group with stenting. Diabetes mellitus and treatment were the predictors that significantly influenced the long-term result in the calculation by logistic regression. Since the study included patients with a satisfactory arterial vessel of the tibia, an analysis of lesion of the tibial vessel as a predictor of restenosis did not reveal a significant effect on long-term results. This study has several limitations, as it was conducted in the format of a single-centre pilot study. There were no complications in operational access in both groups. These methods of treatment are comparable in terms of safety. The method of lamina vastoadductoria dissection after SFA lesion stenting (TASC-II D) showed good primary patency in 2 years. The frequency and severity of stent fractures were significantly lower in the second group. Improvement of the ABI index and clinical parameters in control points, such as the degree of limb ischemia, were also observed. However, further studies are required to evaluate the results of using this method with the participation of more patients, and it is also probably worth excluding from further research patients with total lesion of the arteries of the leg and diabetes mellitus.

## Conclusions

- Lamina vastoadductoria dissection after superficial femoral artery stenting is safe
- The change in biomechanical properties improves primary patency rates in stenting
- Lamina vastoadductoria dissection reduces frequency/severity of stent fractures
- The new method has few early and mid-term postoperative complications

## Data Availability

All data produced in the present work are contained in the manuscript

## Acknowledgements

Special thanks to the Elsevier Language Editing Services for translating the article.

## Notes

### Competing Interest Statement

The authors have declared no competing interest.

### Clinical Trial

NCT02590471

### Funding Statement

This study did not receive any funding

### Author Declarations

This pilot randomised prospective single-centre study was approved by the Local Ethics Committee of Meshalkin National Medical Research Centre, Ministry of Public Health of the Russian Federation (September 15, 2015, protocol No. 53)

